# Variation in the Availability and Cost of Essential Medicines for Non-Communicable Diseases in Uganda: a descriptive time series analysis

**DOI:** 10.1101/2020.02.21.20026393

**Authors:** Mari Armstrong-Hough, Srish Sharma, Sandeep P. Kishore, Ann R Akiteng, Jeremy I. Schwartz

## Abstract

**Background:** Availability of essential medicines for non-communicable diseases (NCDs) is poor in low- and middle-income countries. Availability and cost are conventionally assessed using cross-sectional data. However, these characteristics may vary over time.

**Methods:** We carried out a prospective, descriptive analysis of the availability and cost of essential medicines in 23 Ugandan health facilities over a five-week period. We surveyed facility pharmacies in-person up to five times, recording availability and cost of 19 essential medicines for NCDs and four essential medicines for communicable diseases.

**Results:** Availability of medicines varied substantially over time, especially among public facilities. Among private-for-profit facilities, the cost of the same medicine varied from week to week. Private-not-for-profit facilities experienced less dramatic fluctuations in price.

**Conclusions:** We conclude that there is a need for standardized, continuous monitoring to better characterize the availability and cost of essential medicines, understand demand for these medicines, and reduce uncertainty for patients.

## 1 Introduction

Essential medicines (EM) are those that are critical to meeting the health needs of a population. The World Health Organization (WHO) calls for these medicines to be accessible—both available and affordable.(1) The WHO Model List of EM provides a global framework, which is adapted by individual countries into their own EM lists.(1,2) Yet the presence of a given medicine on a national list does not ensure access.(3-6) High or inconsistent cost, and low or inconsistent availability, are important barriers to access.

For patients with chronic conditions, such as non-communicable diseases (NCDs), consistent access to medicines is critical and represents specific priority areas of the WHO Global Monitoring Framework for NCDs(7) and the Sustainable Development Goals. (8) However, availability and cost may vary across time.(9) Moreover, in some low- and middle-income countries (LMIC), such as Uganda, variation in availability and cost may be greater among EM for NCDs than EM for communicable diseases (CD) because common CD medicines are supplied to facilities via a “push” system, while NCD medicines are supplied via a “pull” system that requires forecasting of facility demand.^10^

We and others have shown that the availability of EM for NCD in LMIC is poor.(3,10) However, the standard methodology for assessing availability and cost of medicines is through cross-sectional, direct observation of a medicine on a pharmacy shelf and documentation of its cost. This strategy ignores variation over time, potentially contributing to inaccurate estimates of availability and cost.(9) Moreover, fluctuations may be more pronounced in LMIC compared to high-income countries.(10)

Approaches that capture the dynamism of EM availability and cost are needed.(9) To explore variation over time, we carried out a prospective, descriptive analysis of the availability and cost of a selection of EM for NCDs in a sample of Ugandan health facilities over a five-week period. We compared this to a selection of EM for acute conditions.

## 2 Methods

### 2.1 Setting

Uganda faces a double burden of CD and NCDs.(11) Even as rates of tuberculosis and HIV/AIDS remain high, the burden of NCDs such as cardiovascular disease and diabetes is increasing and expected to grow rapidly.(12-15) In Uganda, healthcare is available in three sectors: public, private-not-for-profit (PNFP), and private-for-profit (PFP). Within these sectors, facilities are arranged in a referral hierarchy, including a community health worker program (Health Center I [HCI]), outpatient facilities (HCII through HCIV), general hospitals (GH) and regional/national referral hospitals.(3) In the public sector, medicines are distributed to pharmacies, located within health facilities, once every eight weeks.(16) If a medicine goes out of stock, it is not resupplied until the next shipment.

Medicines at public pharmacies are free of charge, while patients pay out of pocket at private-sector pharmacies.

### 2.2 Sample and data collection

We sampled 23 health facilities representing public, PNFP, and PFP sectors from 209 facilities that had participated in the 2013 Service Availability and Readiness Assessment (SARA).(3) We excluded facilities >100km from Kampala, where the population of Uganda is most concentrated. We surveyed facility pharmacies in-person up to five times over a five-week period between 29 August 2016 and 26 September 2016. We recorded availability and cost of the 19 medicines for diabetes, hypertension, chronic lung disease, and mental health in the Essential Medicines and Supplies for Uganda List 2012(17), the current version at the time of data collection: metformin, glibenclamide, soluble insulin, isophane insulin, Mixtard insulin, nifedipine, atenolol, propranolol, captopril, enalapril, lisinopril, losartan, simvastatin, beclomethasone inhaler, salbutamol inhaler, salbutamol tablets, fluoxetine, amitriptyline, and diazepam. We collected the same data for four EM used in the management of acute, infectious conditions: paracetamol, amoxicillin, artemether/lumefantrine, and oral rehydration salts. Availability was documented dichotomously as in stock or out of stock. Cost of each medicine was documented per tablet or vial. Cost was not standardized to course of treatment in any way, as treatment may vary by individual patient. We collected data on facility type, sector, and urban/rural status.

### 2.3 Analysis

This study was designed for a descriptive analysis to support hypothesis generation. The primary unit of analysis was the presence of medicine *in a given facility*. From a patient-centered perspective, the availability of a medicine at their health facility of choice is most important. Therefore, the trajectory of availability over time at a given facility and correlation within a facility of availability and cost are informative metrics.

We defined availability as the proportion of listed EM that were present at a site; if a given medicine was present in any quantity, it was considered available. We calculated the proportion of observations during which each individual EM was available by sector. We plotted the trajectory of medicine availability for each facility with three or more observations using a time series. We tested the hypothesis that week-to-week variations were greater among public sector clinics than private sector facilities using t-tests of change in availability during each period. We tested the hypothesis that there is greater variation in availability of EM for NCD than EM for acute conditions by comparing the intra-cluster correlation coefficient (ICC) of EM availability by facility over time for these two categories of medicines.

We defined variation in cost as the range of fluctuation in price per tablet or vial in a given facility during the study period. We calculated and produced visualizations of these within-facility ranges. We also calculated the quantile-based robust standard deviation of medicine price over time by facility, a conventional measure used to summarize variation in a set of values.

## 3 Results

We collected 82 observations from 23 unique facilities over a five-week period. Each facility had between one and five observations; 21 (91%) facilities contributed two or more observations and the median number of observations was four.

### 3.1 Facility characteristics

We collected observations from five HC-II (22%), nine HC-III (39%), three HC-IV (13%), five GH (22%), and one referral hospital (4%) (Table 1). Twenty-six percent were public, 35% PNFP, and 39% PFP. The majority (83%) of facilities were located in an urban area.

**Table 1.** Characteristics of facilities

| **Facility type** | **N (%)** |
| --- | --- |
| HC-II | 5 (21.7) |
| HC-III | 9 (39.1) |
| HC-IV | 3 (13.0) |
| General Hospital | 5 (21.7) |
| Referral | 1 (4.4) |
| **Managing Authority** |  |
| Public | 6 (26.1) |
| Private-non-profit | 8 (34.8) |
| Private-for-profit | 9 (39.1) |
| **Location** |  |
| Urban | 19 (82.6) |
| Rural | 4 (17.4) |

### 3.2 Variation in medicine availability and cost

EM for NCDs were available at 44% of all study visits, while EM for acute conditions were available at 93% of visits. The proportion of visits in which a medicine was present varied by medicine and by sector (Figure 1). Presented as a time series, the proportion of EM available fluctuated within clinics over the five-week period of observation (Figure 2). Variation in availability over time within the same facility was not significantly different among EM for NCDs (intra-facility correlation coefficient (ICC) 0.68, 95% CI 0.49-0.83) and EM for acute conditions (ICC 0.20, 95% CI 0.04-0.61).

**Figure 1.**
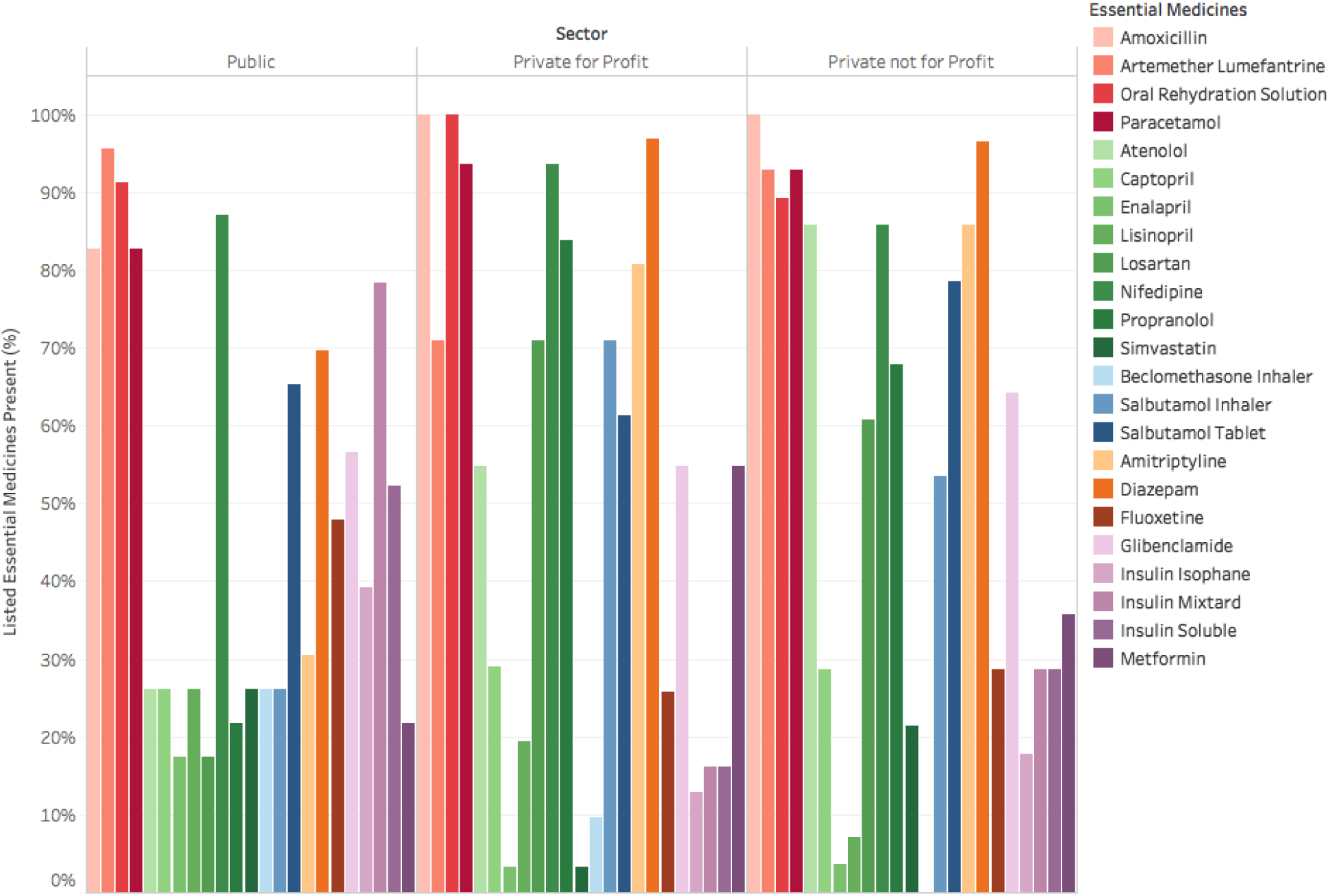
Proportion of visits in which essential medicines were present, by sector. **Legend**: Proportion of visits in which each listed essential medicine was available is depicted by the colored bars and compared by sector (Public, Not for Profit, and Private Not for Profit). The color of each bar corresponds to a specific essential medicine, depicted in the key to the right of the graph.

**Figure 2.**
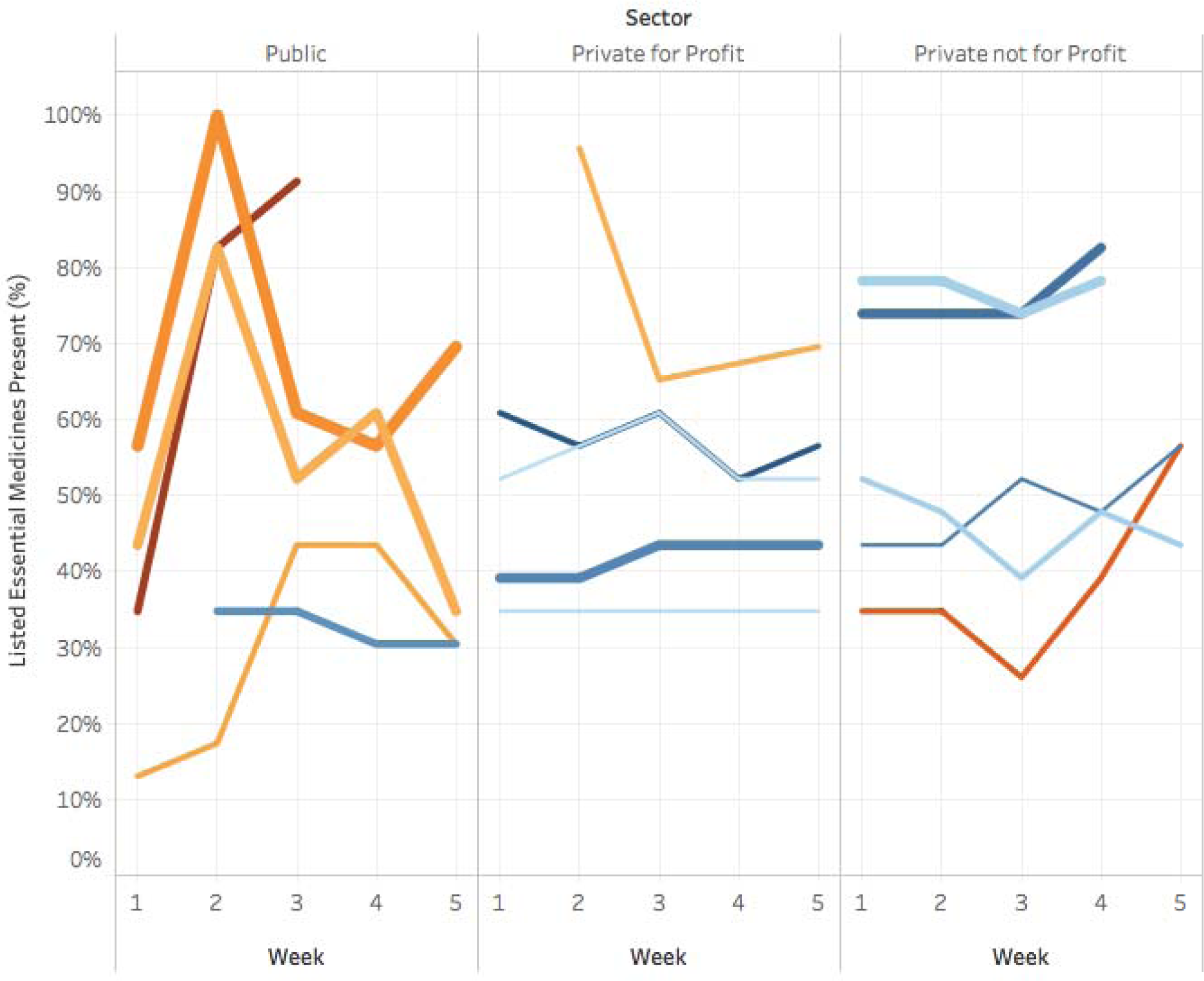
Change in availability of essential medicines over 5 weeks, by sector. **Legend**: These time series plots show the week-by-week shifts in the proportion of essential medicines available by facility, broken down by sector. Each colored line represents one facility; movement on the y axis from week to week indicates change in the proportion of listed essential medicines available in the facility.

Fluctuations in availability were significantly greater among public sector facilities than among PNFP or PFP facilities between Week 1 and Week 2 (p=0.0002), but not during other periods. This is visually evident in Figure 2; public sector facilities show sharp increases and decreases in number of medicines available. There was no significant difference between public and private sector facilities for change in availability during the subsequent periods. This may be attributable to missing data in some subsequent periods: up to 10 missing observations for each period, or 43% of the total sample of 23 facilities. Alternatively, this may be attributable to the periodicity of public sector resupply.

More than half of medicines experienced major price fluctuations, which we defined as at least doubling in price one or more times over the five-week study period (Figure 3). Variation over five weeks in cost to patients was lower in PNFP facilities, with robust standard deviations ranging from UGX 50-704 (approximately US$0.01-$0.19). Among PFP facilities, robust standard deviations in cost of available medicines ranged from UGX 37-17,050 (approximately US$0.01-$4.55).

**Figure 3.**
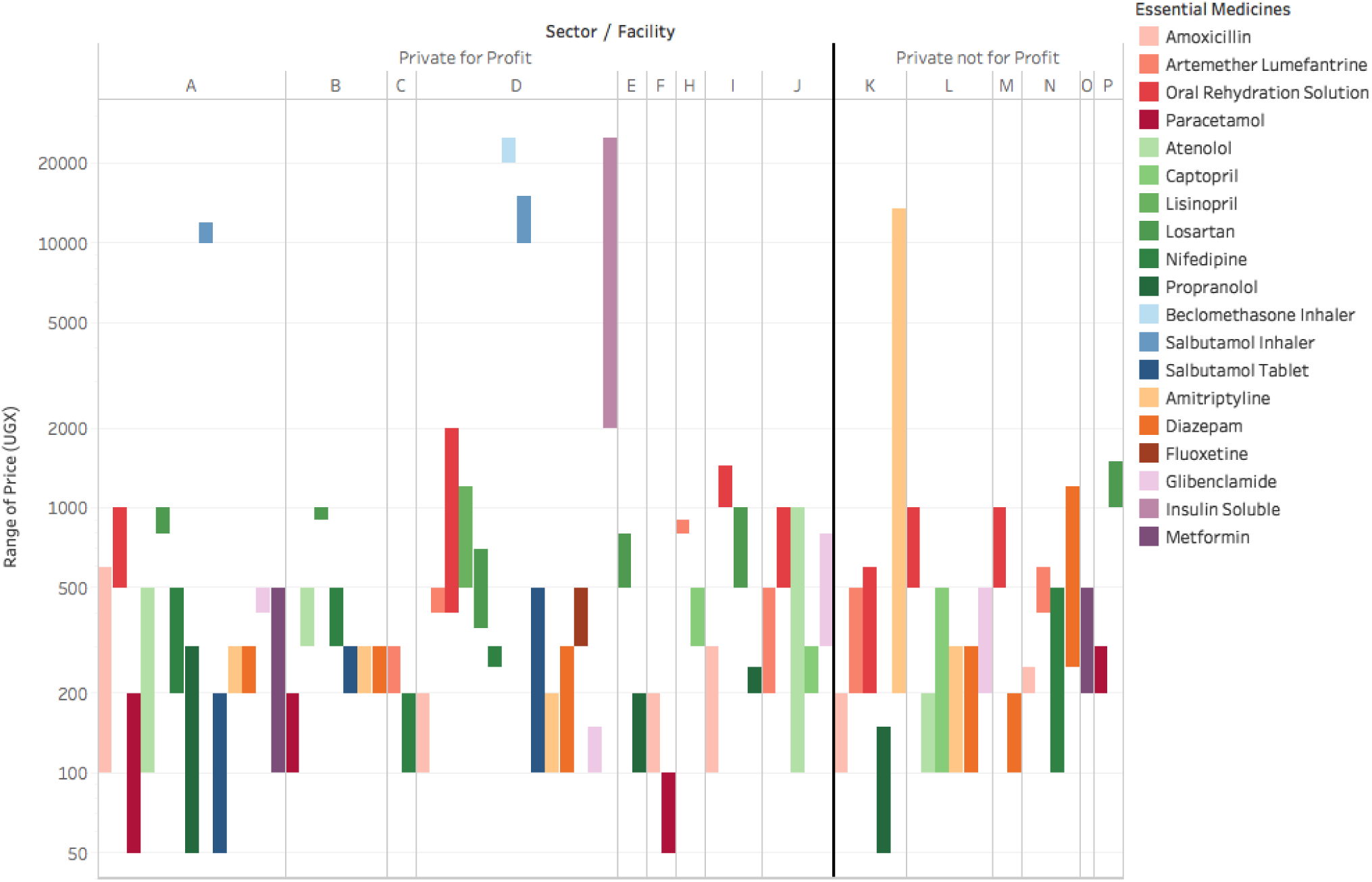
Range of cost over time by clinic and sector. **Legend**: Each bar represents the range in cost of a single medicine within a single clinic over the five-week observation period.

## 4 Discussion

We found that variation in availability was greater among public sector facilities than among PFP and PNFP facilities. In the public sector facilities, more than half of EM alternated in and out of stock during the five-week observation period. We also found that week-to-week fluctuations in the cost of EM were substantial and that PFP facilities had the greatest variations in cost. While PFP facilities are the most likely to have EM for NCD in stock, substantial changes in price from week to week introduce uncertainty for patients and can limit access.

Variations in availability and cost have implications for patient engagement in care. Most medicines for NCDs must be taken regularly over long periods of time. Even in settings where medicines are consistently available, patient adherence to prescription medicines for chronic conditions can be challenging to establish and sustain.(18-20) Inconsistent availability significantly impacts patient adherence to NCD medicines.(21,22) We propose that uncertainty regarding the week-to-week availability or cost of medicines is likely to impair initiation or continuation of treatment, patients’ motivation to adhere to treatment, and patients’ engagement in care. This is especially the case for patients who face other access barriers such as distrust of the medical system, transportation costs, and competing time commitments.

Such week-to-week variations in availability and cost are not captured by the cross-sectional methods typically used to assess access to EM. We have previously shown that cross-sectional assessments can be used to examine disparities in availability of EM.(3) However, given that Ugandan public sector facilities are supplied in bimonthly cycles, the availability of EM can range dramatically. As time since the most recent shipment increases, the number of stocked EM declines and patients may need to shop several facilities, including private sector facilities, to secure medicines—or go without.

Together, these findings point to a need for supply chain management systems that track medicine availability and cost in real-time. To maximize transparency for health system planners, such systems should be incorporated into national health management information systems, alongside epidemiologic surveillance data to guide evidence-based distribution of resources. Allowing citizens to access these data in a user-friendly format would provide an additional layer of transparency, facilitating informed decisions about their care-seeking behavior.

This study had some limitations. First, we collected data for only a five-week period, so we were unable to examine the effect of seasonality or cyclical delivery schedules on availability. Second, the relatively small sample size, nonrandom sampling strategy, and partially missing data limited our time series analysis to description and was not sufficient to support a multivariable model. However, to our knowledge, this is the first published analysis to prospectively describe the dynamism of availability and cost of EM in a LMIC. We also demonstrate how dynamic data on availability and cost of medicines can be collected and analyzed.

## 5 Conclusion

Availability and cost of EM in Uganda can vary sharply within the same facility from week to week. There is a need for standardized, continuous monitoring to better characterize the availability and cost of EM, understand demand for these medicines, and reduce uncertainty for patients by stabilizing availability and cost.

## Data Availability

The raw data supporting the conclusions of this manuscript will be made available by the authors, without undue reservation, to any qualified researcher.

## Abbreviations

UGX: Ugandan Shillings.

## Conflict of Interest

*The authors declare that the research was conducted in the absence of any commercial or financial relationships that could be construed as a potential conflict of interest*.

## Funding

The MacMillan Center of Yale University and the Yale Global Health Leadership Institute provided funding to support travel for JIS through the Hecht-Albert Pilot Innovation Award for Junior Faculty. Knoema provided funding for data collection and development of data collection and management tool.

## Notes

### Competing Interest Statement

The authors have declared no competing interest.

## REFERENCES

1. World Health Organization. The Selection and Use of Essential Medicines. World Health Organization; 2016. 1 p.

2. Kishore SP, Blank E, Heller DJ, Patel A, Peters A, Price M, et al. Modernizing the World Health Organization List of Essential Medicines for Preventing and Controlling Cardiovascular Diseases. J Am Coll Cardiol. 2018 Feb 6;71(5):564–74.

3. Armstrong-Hough M, Kishore SP, Byakika S, Mutungi G, Nunez-Smith M, Schwartz JI. Disparities in availability of essential medicines to treat non-communicable diseases in Uganda: A Poisson analysis using the Service Availability and Readiness Assessment. Khan HTA, editor. PloS one. Public Library of Science; 2018;13(2):e0192332.

4. Hogerzeil HV, Liberman J, Wirtz VJ, Kishore SP, Selvaraj S, Kiddell-Monroe R, et al. Promotion of access to essential medicines for non-communicable diseases: practical implications of the UN political declaration. Lancet. 2013 Feb 23;381(9867):680–9.

5. Kishore SP, Kolappa K, Jarvis JD, Park PH, Belt R, Balasubramaniam T, et al. Overcoming Obstacles To Enable Access To Medicines For Noncommunicable Diseases In Poor Countries. Health Aff (Millwood). Project HOPE - The People-to-People Health Foundation, Inc; 2015 Sep;34(9):1569–77.

6. Siddharthan T, Ramaiya K, Yonga G, Mutungi GN, Rabin TL, List JM, et al. Noncommunicable Diseases In East Africa: Assessing The Gaps In Care And Identifying Opportunities For Improvement. Health Aff (Millwood). Project HOPE - The People-to-People Health Foundation, Inc; 2015 Sep;34(9):1506–13.

7. World Health Organization. Noncommunicable Diseases Global Monitoring Framework: Indicator Definitions and Specifications. Geneva: World Health Organization; 2014.

8. United Nations. The Sustainable Development Goals Report 2018. New York; 2018.

9. Robertson J, Macé C, Forte G, de Joncheere K, Beran D. Medicines availability for non-communicable diseases: the case for standardized monitoring. Global Health. BioMed Central; 2015 May 7;11(1):18.

10. Yadav P, Stapleton O, Van Wassenhove LN. Always cola, rarely essential medicines: comparing medicine and consumer product supply chains in the developing world. 2010.

11. World Health Organization. Uganda: WHO statistical profile. 2015.

12. Chiwanga FS, Njelekela MA, Diamond MB, Bajunirwe F, Guwatudde D, Nankya-Mutyoba J, et al. Urban and rural prevalence of diabetes and pre-diabetes and risk factors associated with diabetes in Tanzania and Uganda. Glob Health Action. 2016;9:31440.

13. Guwatudde D, Mutungi G, Wesonga R, Kajjura R. The epidemiology of hypertension in Uganda: findings from the national non-communicable diseases risk factor survey. Kokubo Y, editor. PloS one. 2015;10(9):e0138991.

14. Schwartz JI, Guwatudde D, Nugent R, Kiiza CM. Looking at non-communicable diseases in Uganda through a local lens: an analysis using locally derived data. Global Health. BioMed Central; 2014 Nov 19;10(1):77.

15. Bahendeka S, Wesonga R, Mutungi G, Muwonge J, Neema S, Guwatudde D. Prevalence and correlates of diabetes mellitus in Uganda: a population-based national survey. Trop Med Int Health. 2016 Mar;21(3):405–16.

16. National Medical Logistics. FY 2017/18 National Medical Stores Delivery Schedule.

17. Ministry of Health ROU. Essential Medicines and Health Supplies List for Uganda [Internet]. apps.who.int. 2012 [cited 2017 Feb 27]. Available from: http://apps.who.int/medicinedocs/en/d/Js21740en/

18. Fung V, Graetz I, Reed M, Jaffe MG. Patient-reported adherence to statin therapy, barriers to adherence, and perceptions of cardiovascular risk. Aalto-Setala K, editor. PloS one. Public Library of Science; 2018;13(2):e0191817.

19. Pittman DG, Chen W, Bowlin SJ, Foody JM. Adherence to statins, subsequent healthcare costs, and cardiovascular hospitalizations. Am J Cardiol. 2011 Jun 1;107(11):1662–6.

20. Rash JA, Campbell DJT, Tonelli M, Campbell TS. A systematic review of interventions to improve adherence to statin medication: What do we know about what works? Prev Med. 2016 Sep;90:155–69.

21. Ali M, Alemu T, Sada O. Medication adherence and its associated factors among diabetic patients at Zewditu Memorial Hospital, Addis Ababa, Ethiopia. BMC Res Notes. 2nd ed. BioMed Central; 2017 Dec 4;10(1):676.

22. Gaziano T, Cho S, Sy S, Pandya A, Levitt NS, Steyn K. Increasing Prescription Length Could Cut Cardiovascular Disease Burden And Produce Savings In South Africa. Health Aff (Millwood). 2015 Sep;34(9):1578–85.

